# Real-world Use of Nirmatrelvir-Ritonavir in COVID-19 Outpatients During the Emergence of Omicron Variants BA.2/BA2.12.1

**DOI:** 10.1101/2022.09.12.22279866

**Authors:** Neil R. Aggarwal, Kyle C. Molina, Laurel E. Beaty, Tellen D. Bennett, Nichole E. Carlson, Adit A. Ginde

## Abstract

**Background:** Ritonavir-boosted Nirmatrelvir (NMV-r), a protease inhibitor with in vitro activity against SARS-CoV-2, has been shown to reduce risk of progression to severe COVID-19 among high-risk individuals during the Delta-variant phase. We sought to determine the effectiveness of NMV-r against Omicron lineage variants BA.2/BA2.12.1, and assess for evidence of a clinical rebound effect.

**Methods:** We conducted a retrospective observational cohort study of non-hospitalized adult patients with SARS-CoV-2 infection from March 26^th^, 2022 to June 23^rd^, 2022, using records from a statewide health system linked to vaccine and mortality data. Propensity score matching was performed on NMV-r treated outpatients with outpatients not treated with antiviral therapy. The primary outcome was 28-day all-cause hospitalization; secondary outcomes were COVID-19-related hospitalization, 28-day all-cause mortality, and 28-day ED visits. Logistic regression was used to determine NMV-r treatment effectiveness; subgroup analyses were performed to assess for heterogeneity in treatment effect.

**Results:** Of 14,953 SARS-CoV-2 infected outpatients, 3,614 NMV-r treated patients were matched to 4,835 untreated outpatients. NMV-r was associated with significantly lower odds of 28-day all-cause hospitalization as compared to no antiviral treatment [31 (0.9%) vs. 64 (1.3%), adjusted odds ratio (aOR): 0.48 (95% CI 0.31-0.75)]. NMV-r was also associated with lower odds of COVID-19 related hospitalization [aOR (95% CI): 0.42 (0.25-0.68)] and 28-day all-cause mortality [aOR (95% CI): 0.05 (0.00-0.38)]. Using ED visits within 28 days as a surrogate for rebound symptoms, we observed no clinically evident rebound effect with NMV-r treatment [140 (3.9%) vs 205 (4.2%), aOR: 0.81 (95% CI 0.65-1.02), p = 0.075].

**Conclusion:** Real-world evidence during an Omicron BA.2/BA2.12.1 predominant period demonstrated an association of NMV-r treatment with reduced 28-day hospitalization and all-cause mortality, and without an increase in rebound symptoms as assessed by ED visits within 28 days after treatment.

## BACKGROUND

The global spread and impact of the severe acute respiratory syndrome coronavirus 2 (SARS-CoV-2) have highlighted the need for accessible therapeutics, which improve patient outcomes and attenuate the effect of COVID-19 surges on the healthcare system. Nirmatrelvir is an orally bioavailable protease inhibitor that has activity against the viral protease main protease (M^PRO^), which is essential to SARS-CoV-2 viral replication.^1^ In the EPIC-HR trial, treatment with ritonavir-boosted nirmatrelvir (NMV-r; Paxlovid; Pfizer Labs, NY) resulted in a risk of progression to severe disease that was 89% lower than placebo among unvaccinated adults during the Delta pandemic phase.^2^ Based on these results, in December 2021, NMV-r was granted U.S. Food and Drug Administration emergency use authorization (EUA) for the treatment of mild-to-moderate coronavirus disease 2019 (COVID-19) in adult and pediatric patients who are at high risk for progression to severe COVID-19, including hospitalization or death.^3^

Since the initial authorization of NMV-r, the landscape of the COVID-19 pandemic has evolved. Omicron lineage variants of SARS-COV-2 that demonstrate high transmissibility and immune evasion, yet are associated with decreased disease severity, have supplanted previous variants.^4^ In addition, early real-world observations have hypothesized the occurrence of an NMV-r rebound effect, whereby treated patients may experience an increase in viral load and/or recurrent symptoms after treatment.^5,6^ Further, vaccination for COVID-19 has become widespread, in comparison, to the unvaccinated population of the EPIC-HR trial. Several non-U.S. based observational studies have demonstrated the benefit of NMV-r treatment primarily during the Delta-variant phase of COVID-19, yet effectiveness data against Omicron variants is limited.^7,8^ Given the epidemiological shift in circulating variants, suggestion of a rebound phenomenon, and extensive vaccination of high-risk individuals, real-world data are critical to evaluate the impact of NMV-r and other therapies targeting COVID-19 to inform ongoing policy and practice decisions. To provide additional data on NMV-r effectiveness against Omicron lineage SARS-CoV-2, we used our real-world data platform to evaluate the impact of NMV-r treatment on hospitalization, ED visits, and mortality among outpatients with early symptomatic COVID-19 during a SARS-CoV-2 Omicron (BA.2/BA2.12.1) predominant phase in Colorado.

## METHODS

### Patient Population

In continuation of prior work, we conducted a propensity-matched observational cohort study, a collaboration between University of Colorado researchers, University of Colorado Health (UCHealth) leaders, and the Colorado Department of Public Health and Environment (CDPHE).^9,10^ The study was approved by the Colorado Multiple Institutional Review Board with a waiver of informed consent. We obtained data from the electronic health record (EHR; Epic, Verona, WI) of UCHealth, the largest health system in Colorado with 13 hospitals around the state and 141,000 annual hospital admissions, using Health Data Compass, an enterprise-wide data warehouse. EHR data were merged with statewide data on vaccination status from the Colorado Comprehensive Immunization Information System and mortality from Colorado Vital Records.

We included all patients diagnosed with SARS-CoV-2 infection as identified using EHR-based date of a SARS-CoV-2 positive test (either polymerase chain reaction or antigen) or date of antiviral order if a SARS-CoV-2 test result was unavailable. Patients were included if their positive test date was between March 26^th^, 2022, and June 23^rd^, 2022, which allowed for a minimum of 28 days of follow-up (n = 14,953) (**Appendix Figure 1, Supplement**). During this period, NMV-r was readily available and the dominant variant was Omicron. The decision to seek antiviral treatment was made by patients and clinicians.^11^ We did not exclude patients who did not meet EUA eligibility criteria based on EHR data, as not all eligibility criteria were consistently available.

The main exclusion criteria were: 1) order or administration of molnupiravir, or adminstration of any other mAb or antiviral [including bebtelovimab, sotrovimab, tixagevimab/cilgavimab (within 10 days of SARS-CoV-2 positive data), or outpatient remdesivir] (n = 3,250), 2) hospitalization at the time of the SARS-CoV-2 positive test or in the hospital at the time of the NMV-r treatment (n = 484), or 3) a positive SARS-CoV-2 test more than 10 days prior to the order date of NMV-r (n = 5). We retained patients who were hospitalized or died on the same day as their observed SARS-CoV-2 positive test, given the common use of home self-testing during the study period. After exclusions, 11,214 patients were available for analysis: 4,672 NMV-r-treated patients and 6,542 untreated patients.

Of note, the majority of patients treated with NMV-r did not have a SARS-CoV-2 positive test date (81.6%) in the health system, which does not capture the results of home testing. Since many patients received oral antiviral treatment the same or next day after a positive SARS-CoV-2 test, we assumed a SARS-CoV-2 positivity test date to be one day before the NMV-r order date for analytic purposes.

To achieve balance on potential confounders, nearest neighbor propensity matching with logistic regression with treatment status as the outcome was used.^12,13^ We achieved a matching ratio of 1.36:1 treated to untreated patients, with a total matched cohort size of 8,449. The propensity model included categorical age, sex, race/ethnicity, insurance status, immunocompromised status, obesity status, number of comorbid conditions other than immunocompromised and obesity, number of vaccinations at the time of infection, and categorical week. We removed 435 NMV-r treated patients due to missing covariate data, and utilized the recommended caliper of 0.2 (**Appendix Table 1, Supplement**).^14^ Variables with a remaining standardized mean difference (SMD) above 0.1 were adjusted for in all models, to account for residual imbalance.^15^

### Outcomes

The primary outcome was all-cause hospitalization within 28 days of a positive SARS-CoV-2 test. We also created a COVID-19 specific 28-day hospitalization defined by the presence of any of the following: COVID-19 ICD-10 codes (U07.1, J12.82, M35.81, Z20.822, M35.89), administration of inpatient remdesivir, or use of any supplemental oxygen. Secondary outcomes for the whole cohort included 28-day all-cause mortality and 28-day all-cause emergency department (ED) visits. In the hospitalized subset, exploratory outcomes included disease severity based on maximum level of respiratory support, hospital and intensive care unit (ICU) lengths of stay (LOS) in survivors, rates of ICU admission, and in-hospital mortality.

### Variable Definitions

Hospitalization was defined as any inpatient or observation encounter documented in the EHR. We selected the first hospitalization that occurred the same day, or any day after, a SARS-CoV-2 positive test for untreated patients, or after the order date for NMV-r for treated patients. ED visits were defined as any visit to the ED, with or without an associated inpatient or observation encounter. For treated patients, we selected the first ED visit that occurred at least one day after the NMV-r order date, given that NMV-r treatment was often prescribed at the initial ED visit (and thus should not be considered a treatment failure outcome). We defined COVID-19 disease severity as the maximum level of respiratory support received in the following order from lowest to highest severity: no supplemental oxygen, standard (nasal cannula/face mask) oxygen, high-flow nasal cannula or non-invasive ventilation, and invasive mechanical ventilation.^16^ In-hospital mortality was the highest level of disease severity.

The variables of interest include treatment status, categorical age in years, sex, race/ethnicity, insurance status, obesity status, immunocompromised status, number of additional comorbid conditions, and number of vaccinations. Presence of comorbid conditions (obesity, hypertension, cardiovascular disease, diabetes mellitus, pulmonary disease, pulmonary disease, and liver disease) was based on the Charlson Comorbidity Index and immunocompromised status was custom coded as reported previously (**Appendix Table 2, Supplement**).^17^ The number of comorbid conditions was calculated as the sum of the presence of hypertension, cardiovascular disease, diabetes, pulmonary disease, pulmonary disease, and liver disease. Obesity and immunocompromised status were kept as separate comorbid conditions in analysis. Vaccination status was categorized by the number of vaccinations (0, 1, 2, or ≥ 3) administered prior to the date of the SARS-CoV-2 positive test.

### Statistical Analysis

We used Firth’s logistic regression to assess the association between treatment and 28-day hospitalization, 28-day mortality, and 28-day ED visits. Firth’s logistic regression (R package logistf V 1.24) addresses estimation issues related to low event rates and complete separation.^15,18,19^ All models were adjusted for age, sex, race/ethnicity, insurance status, obesity status, immunocompromised status, number of additional comorbid conditions, number of vaccinations, and week in the study. Kaplan-Meier curves were estimated to visually assess the time from treatment to ED visit in those who received an order for NMV-r. Care should be used in interpreting these curves due to use of rapid antigen home testing prior to a health care encounter for PCR or treatment. Due to the small number of hospitalized participants, we present only descriptive statistics for all exploratory outcomes among hospitalized participants, including disease severity, hospital LOS, ICU visit, and ICU LOS.

We estimated adjusted treatment effects for four subgroups of interest by fitting interaction models that were also adjusted for all variables of interest. The subgroups of interest included binary age (<65 vs. ≥65), immunocompromised status (both binary, and three levels), binary number of comorbidities (0-1 vs. ≥2), and binary vaccination status (0-2 vs. ≥3). All statistical analyses were performed using R Statistical Software (version 3.6.0; R Foundation for Statistical Computing, Vienna, Austria).

## RESULTS

### Characteristics of NMV-r treated and untreated cohorts in the primary cohort

Among 14,953 patients with positive SARS-CoV-2 test in the study window, 11,214 patients met study inclusion, and a total of 4,672 patients received treatment with NMV-r and 6,542 were untreated (**Appendix Figure 1**). Propensity-score matching resulted in 8,449 patients (3,614 NMV-r, 4,835 Untreated) for the primary analysis, with balance observed between most key prognostic factors (**Appendix Table 3**).

The characteristics of the propensity matched primary cohort are presented in **Table 1**. The NMV-r cohort generally reflect characteristics of patients at high risk for progression to severe COVID-19. Among the NMV-r treated patients, 34.9% were ≥65 years of age (34.9%), 27.8% were obese, 26.0% were immunocompromised, and 32.3% had two or more comorbid conditions.

**Table 1:** Baseline Characteristics by NMV-r Treatment Status for Primary Matched Cohort.

|  | **NMV-r**<br>**(N=3,614)** | **Untreated**<br>**(N=4,835)** |
| --- | --- | --- |
| **Age Group\*** |  |  |
| 18-44 years | 1,596 (44.2%) | 2,991 (61.9%) |
| 45-64 years | 755 (20.9%) | 774 (16.0%) |
| ≥65 years | 1,263 (34.9%) | 1,070 (22.1%) |
| **Female Sex** | 2,133 (59.0%) | 2,798 (57.9%) |
| **Race/Ethnicity** |  |  |
| Non-Hispanic White | 3,006 (83.2%) | 3,929 (81.3%) |
| Hispanic | 351 (9.7%) | 477 (9.9%) |
| Non-Hispanic Black | 115 (3.2%) | 222 (4.6%) |
| Other | 142 (3.9%) | 207 (4.3%) |
| **Insurance Status\*** |  |  |
| Private/Commercial | 2,187 (60.5%) | 3,367 (69.6%) |
| Medicare | 1,166 (32.3%) | 1,033 (21.4%) |
| Medicaid | 155 (4.3%) | 267 (5.5%) |
| None/Uninsured | 18 (0.5%) | 54 (1.1%) |
| Other/Unknown | 88 (2.4%) | 114 (2.4%) |
| **Immunocompromised\*** | 940 (26.0%) | 824 (17.0%) |
| Mild | 454 (12.6%) | 397 (8.2%) |
| Moderate/Severe | 486 (13.4%) | 427 (8.8%) |
| **Obese\*** | 1,003 (27.8%) | 974 (20.1%) |
| **Number of Other Comorbid Conditions** |  |  |
| One | 1,073 (29.7%) | 1,365 (28.2%) |
| Two or more | 1,167 (32.3%) | 1,038 (21.5%) |
| **Diabetes Mellitus** | 555 (15.4%) | 472 (9.8%) |
| **Cardiovascular Disease** | 584 (16.2%) | 600 (12.4%) |
| **Pulmonary Disease** | 1,063 (29.4%) | 1,111 (23.0%) |
| **Renal Disease** | 246 (6.8%) | 261 (5.4%) |
| **Hypertension** | 1,377 (38.1%) | 1,335 (27.6%) |
| **Liver Disease** | 332 (9.1%) | 330 (6.8) |
| Mild | 318 (8.8%) | 307 (6.3%) |
| Severe | 14 (0.4%) | 23 (0.5%) |
| **Number of vaccinations prior to SARS-CoV-2+ date\*** |  |  |
| 0 | 654 (18.1%) | 953 (19.7%) |
| 1 | 142 (3.9%) | 201 (4.2%) |
| 2 | 488 (13.5%) | 732 (15.1%) |
| 3+ | 2,330 (64.5%) | 2,949 (61.0%) |

\* Variables used in the propensity matching. Abbreviations: NMV/r Nirmatrelvir-ritonavir

### Primary Outcome

Treatment with NMV-r was associated with significantly lower odds of 28-day all cause hospitalization as compared to no antiviral treatment [31 (0.9%) vs. 64 (1.3%), adjusted odds ratio (aOR): 0.48 (95% CI 0.31-0.75), p < 0.001] (**Table 2, Figure 1**). Furthermore, COVID-related hospitalizations constituted approximately 80% of all hospitalizations, and among this subset the odds of 28-day hospitalization among NMV-r treated were reduced to a similar level compared to no antiviral treatment [23 (0.6%) vs. 52 (1.1%), aOR (95% CI): 0.42 (0.25-0.68), p < 0.001].

**Table 2:** Primary and Secondary Outcomes for NMV-r Treatment for Primary Matched Cohort.

|  | **NMV-r**<br>**(N = 3,614)** | **Untreated**<br>**(N = 4,835)** | **Adjusted OR**<br>**(95% CI)** | **P-Value** |
| --- | --- | --- | --- | --- |
| All-Cause 28-day Hospitalization | 31 (0.9%) | 64 (1.3%) | 0.48 (0.31-0.75) | <0.001 |
| COVID-related 28-day Hospitalization<sup>a</sup> | 23 (0.7%) | 52 (1.1%) | 0.42 (0.25-0.68) | <0.001 |
| All-cause 28-day ED visit | 140 (3.9%) | 205 (4.2%) | 0.81 (0.65-1.02) | 0.075 |
| All-cause 28-day Mortality | 0 (0.0%) | 9 (0.2%) | 0.05 (0.00-0.38) | 0.001 |
| Hospital LOS, days, Mean (SD) | 3.9 (5.2) | 4.9 (5.4) | -- | -- |
| ICU Visit During Hospitalization<sup>b</sup> | 2 (6.5%) | 10 (15.6%) | -- | -- |

All regression models adjusted for age, sex, race/ethnicity, obesity, immunocompromised status, number of comorbidities, insurance status, and vaccination status Abbreviations: NMV-r, Nirmatrelvir-ritonavir; OR, odds ratio; CI, Confidence Interval; LOS, Length of Stay; ICU, Intensive Care Unit.
- a. COVID-related hospitalization was defined by presence of COVID-19 ICD-10 codes (U07.1, J12.82, M35.81, Z20.822, M35.89), administration of inpatient remdesivir, or use of supplemental oxygen - b. Among 31 and 64 all-cause hospitalizations in the NMV/r and Untreated groups, respectively.

**Figure 1.**
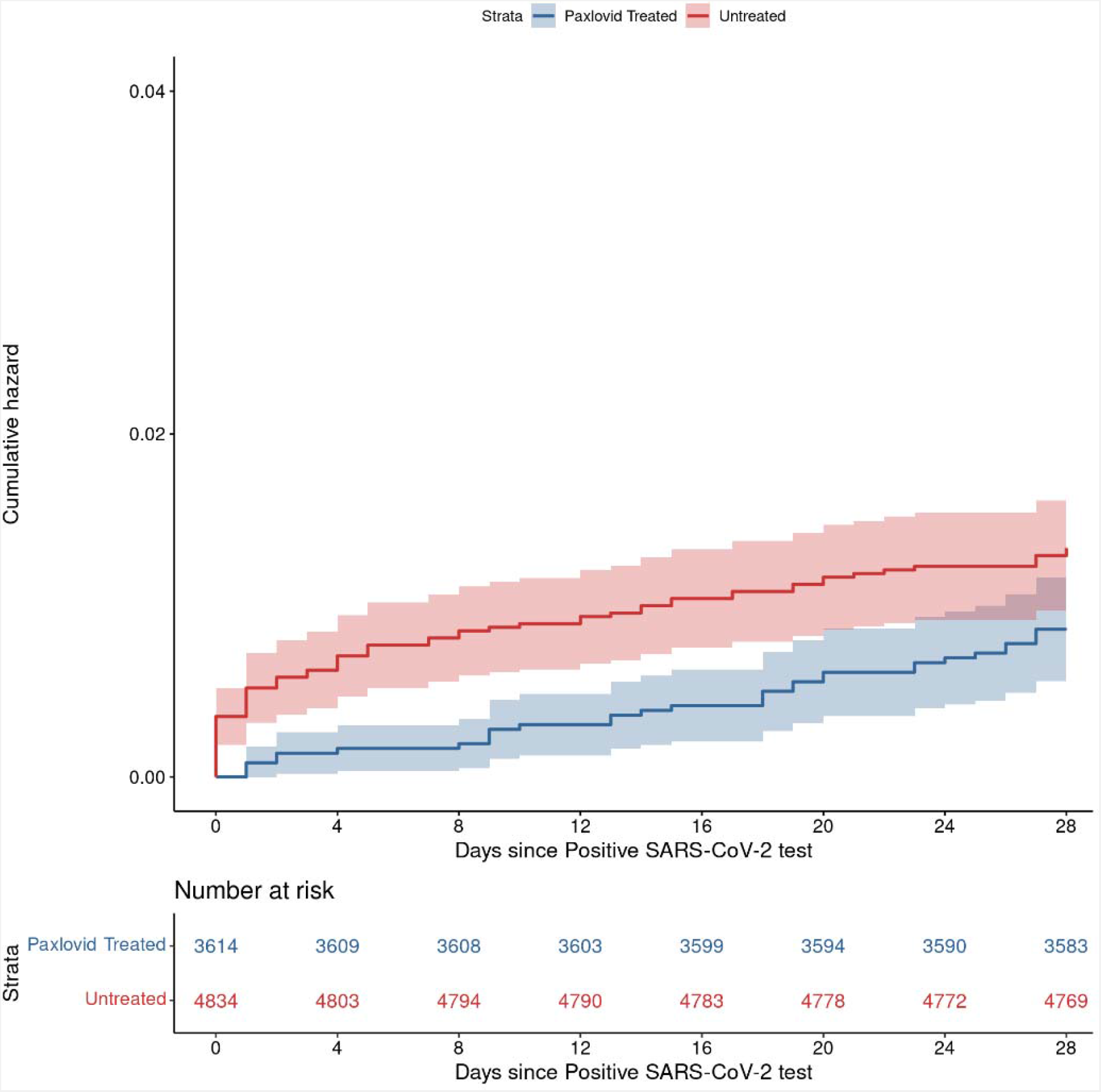
Cumulative Incidence Plots for All-Cause Hospitalization to Day 28 by NMV-r Treatment Status.

### Secondary Outcomes

28-day all-cause mortality, a key indicator of severity of illness, was significantly lower among those treated with NMV-r compared to untreated patients [0 (0.0%) vs. 9 (0.2%), aOR (95% CI): 0.05 (0.00-0.38), p = 0.001] (**Table 2**). Furthermore, among the subset of hospitalized patients in our cohort, the need for high flow nasal oxygen, invasive mechanical ventilation, or death was lower in NMV-r treated patients compared to untreated patients (6.4% vs. 10.9%), though inferential statistics could not be performed due to low event rates (**Figure 2**).

**Figure 2.**
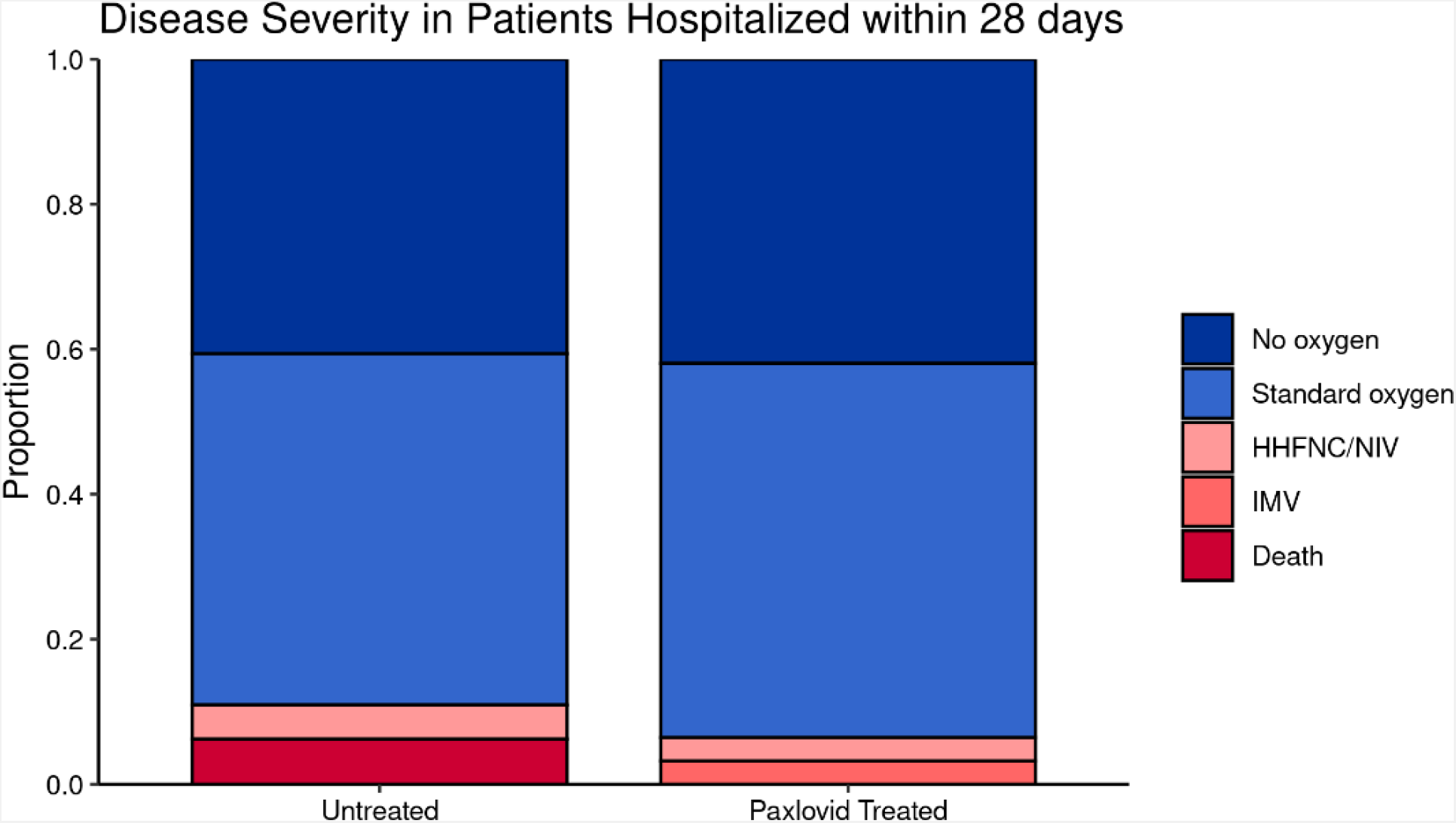
Severity of All-Cause Hospitalization Through Day 28. The total sample size of the hospitazled subset is 95, of which 31 and 64 are in the Nirmatrelvir-ritonavir and untreated group, respectively.

We used ED visits as a surrogate for severe symptoms to evaluate for a rebound effect after NMV-r, because we cannot track COVID-related symptoms in our EHR-derived cohort. The time to ED visit from positive SARS-CoV-2 test in each cohort is displayed in **Figure 3**, which notably does not demonstrate a visual discontinuity (or jump) after NMV-r treatment completion that may be anticipated with a severe rebound effect after treatment. Overall, the NMV-r treated group had similar odds of ED visit within 28 days compared to untreated controls [140 (3.9%) vs 205 (4.2%), aOR: 0.81 (95% CI 0.65-1.02), p = 0.075] (**Table 2**).

**Figure 3:**
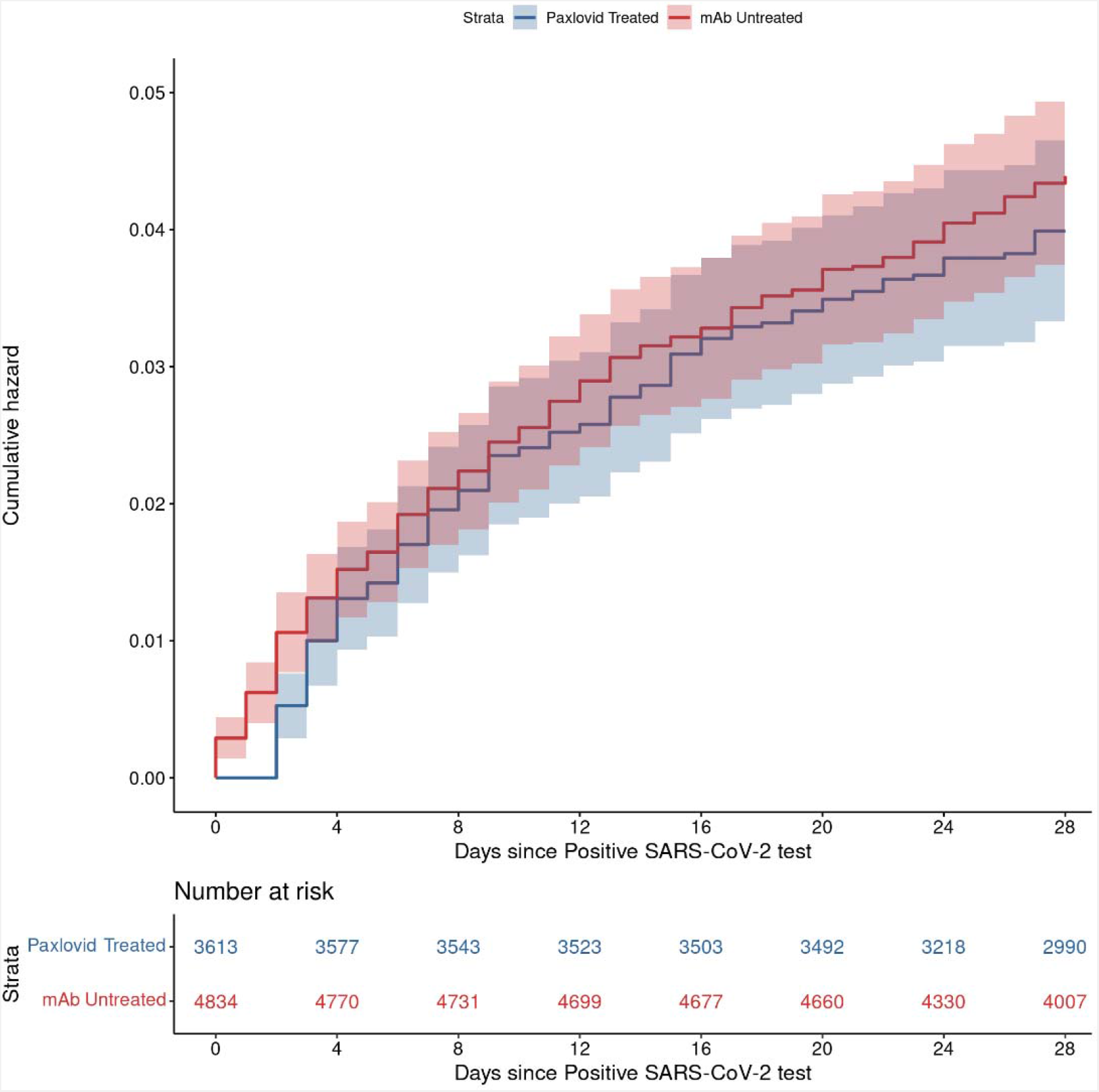
Cumulative Incidence Plots for All-Cause Emergency Department Visit to Day 28 by NMV-r Treatment Status.

Important subgroups based on age, immunocompromised status, vaccine status, or number of comorbidities were also evaluated (**Table 3**). Treatment effect differed only based on the number of comorbid conditions; although statistical significance was marginal (Table 3; p=0.059). In those with 2+ comorbid conditions other than immunocompromised status or obesity, there was a significant reduction in the odds of 28-day all-cause hospitalization [42 (4.1%) vs. 18 (1.5%), aOR: 0.35 (95% CI: 0.2-0.62), p<0.001]. There was no effect of treatment on 28-day all-cause hospitalization in those with 0 or 1 comorbid conditions [22 (0.6%) vs 13(0.5%), aOR: 0.84 (95% CI: 0.41-1.65),p=0.61]. Treatment effects were similar for different ages (p=0.99), immunocompromised status (p=0.79), and vaccine status (p=0.69).

**Table 3:** Subgroup Analysis for 28-day All-Cause Hospitalization.

|  |  | **N (%) Hospitalized** |  |  |  |
| --- | --- | --- | --- | --- | --- |
|  | **N** | **NMV-r** | **Untreated** | **OR (95% CI)** | **P-value** |
| **Age** |  |  |  |  | **0.99** |
| < 65 years | 6,116 | 13 (0.55%) | 35 (0.93%) | 0.47 (0.24-0.87) | 0.022 |
| ≥ 65 years | 2,333 | 18 (1.43%) | 29 (2.71%) | 0.47 (0.26-0.86) | 0.015 |
| **Immunocompromised** |  |  |  |  | **0.79** |
| No | 6,685 | 15 (0.56%) | 36 (0.9%) | 0.51 (0.27-0.92) | 0.03 |
| Yes | 1,764 | 16 (1.7%) | 28 (3.4%) | 0.45 (0.24-0.84) | 0.013 |
| **Number of Comorbidities** |  |  |  |  | **0.059** |
| 0-1 | 6,244 | 13 (0.53%) | 22 (0.58%) | 0.84 (0.41-1.65) | 0.613 |
| 2+ | 2,205 | 18 (1.54%) | 42 (4.05%) | 0.35 (0.2-0.62) | <0.001 |
| **Number of Vaccinations** |  |  |  |  | **0.69** |
| 0-2 | 3,170 | 15 (1.17%) | 33 (1.75%) | 0.53 (0.27-0.97) | 0.044 |
| 3+ | 5,279 | 16 (0.69%) | 31 (1.05%) | 0.44 (0.24-0.81) | 0.009 |

## DISCUSSION

During a SARS-CoV-2 Omicron (BA.2/BA2.12.1) predominant phase in a US state, NMV-r was associated with a lower incidence of the primary outcome of 28-day all-cause hospitalization, and associated with a lower incidence of 28-day all-cause hospitalization using a more restrictive definition of COVID-related hospitalization. NMV-r administration to high-risk outpatients was also associated with a reduction in COVID illness severity, as evidenced by a reduction in all-cause 28-day mortality, and suggestion of reduced levels of maximal respiratory support, ICU needs, or hospital length of stay. Therefore, despite waning COVID severity and increasing numbers of non-specific hospitalizations among COVID-positive patients, we observed a strong association of benefit for NMV-r against SARS-CoV-2 Omicron during a BA.2/BA2.12.1 phase.

The other major finding of this study was that NMV-r was not associated with an increase in ED visits in the 28 days following its administration, and particularly in the 10-14 days after anticipated completion of the treatment course, as compared to matched, untreated control patients. These data suggest that patients who are treated with NMV-r may not develop severe rebound symptoms that lead to ED evaluation with greater frequency than untreated individuals. Systematically tracking all symptoms and viral testing in treated and untreated COVID-19 patients would be ideal; however, that is not feasible in our real-world setting, and we elected to use ED visits as a surrogate for illness severity during the post-treatment phase. Our findings support the study by Malden et al who found that ED visits or hospitalizations occurred with less than one percent frequency in the 5-15 days after NMV-r treatment.^6^

Our findings also support observed NMV-r neutralization of Omicron variants in vitro and those of recently published data in high-risk outpatients and lower severity inpatients.^7,8,20^ Notably, the study by Arbel et al. found that high-risk outpatients in Israel aged 40-64 and infected with COVID-19 did not appear to derive benefit from nirmaltrelvir with respect to reduced hospitalization, yet among patients 65 or older, the adjusted HR of 0.21 suggested striking benefit from nirmaltrelvir.^7^ In contrast, we observed that NMV-r may have been beneficial in both age groups. This discrepancy is not readily explained by differences in methodology or in risk factors for COVID severity in the respective cohorts, and may instead be due to differences in setting, including thresholds for hospitalization in younger patients, population differences, or other unmeasured factors.

### Limitations

This study has several limitations. Hospitalizations were collected only within a single health system that has relatively low racial and ethnic minority representation despite providing care in both urban and rural settings at academic and community hospitals. If untreated patients were more likely to be hospitalized outside this health system, this may bias our results toward the null.

SARS-CoV-2 test result missingness was high and unbalanced in this cohort, with more than 80% of NMV-r treated patients without test results in our system. At the present juncture in the pandemic, we speculate this missingness was so high because symptomatic people were testing at home and reporting those results to providers who then prescribed NMV-r. As such, we imputed the test date based on the NMV-r order date and knowledge of clinical practice patterns for those in the NMV-r treated group. Unlike in other analyses we did not exclude hospitalizations on the date of a positive test.^10^ This was again owing to changes in testing. These approaches may introduce bias in the early days of the time to event analysis, and as such, the Kaplan-Meier results should be interpreted with caution.

Finally, NMV-r treatment was assigned based on NMV-r order, however, NMV-r treatment was not directly observed. Yet, this would likely bias comparisons against a beneficial effect of NMV-r as some patients may have received partial or no treatment.

## Conclusion

This study of real-world data demonstrated NMV-r treatment was associated with reduced 28-day hospitalization and all-cause 28-day mortality among high-risk COVID-19 outpatients during the Omicron BA.2/BA2.12.1 variant phase in Colorado. Using ED visits as a surrogate for severe and/or recurrent symptoms during the treatment phase, we did not observe a higher ED visit rate in NMV-r treated patients as compared to untreated patients, and thus cannot conclude that treatment with NMV-r induces severe rebound symptoms.

## Data Availability

All data produced in the present study are available upon reasonable request to the authors

