## Supplementary Material for "Real-world Use of Nirmatrelvir-Ritonavir in COVID-19 Outpatients During the Emergence of Omicron Variants BA.2/BA2.12.1"

This Appendix has been provided by the authors to give readers additional information about the work.

**Appendix Figures**

**Appendix Figure 1: Flow of Patients into the Primary Study Cohort**

Hospitalization Criteria: Removed N = 484

- Subject was already admitted to the hospital at the time of their COVID + test (N = 476)
  - Note: 2 treated patients were removed
- Subject was currently in the hospital at the time of the Paxlovid order (N = 2)
- Subject was discharged the same date as the Paxlovid order (N = 6)

Propensity Matching

N = 14,928 Selecting only patients that are missing a covid+ date

missing_covid_date_df <- current_df[is.na(current_df$covid_pos_dttm),]

dim(missing_covid_date_df) # Checking to make sure that we have all of the patients that we # Selecting only patients that are missing a covid+ date

missing_covid_date_df <- current_df[is.na(current_df$covid_pos_dttm),]

dim(missing_covid_date_df) # Checking to make sure that we have all of the patients that we need

N = 14,953

N = 11,703

Subject had a positive test more than 10 days prior to the order date for Paxlovid (N = 5)

N Untreated = 4,835

N Treated = 3,614

Subsetting to untreated and Paxlovid treated only

- Removed: treated with Evusheld within 10 days of the positive test (N = 541), bebtelovimab (N = 2,152), sotrovimab (N = 121), outpatient remdesivir (408), or an administration of Paxlovid without an order (3)

103,922 subjects with a SARS CoV 2 positive test date on or before 6/23/22

Subject had a positive test prior to Paxlovid being readily available (before 3/26/2022): N = 88,969

N = 11,219

Subject had an order for molnupiravir (N = 25)

N = 11,214

**Appendix Tables**

**Appendix Table 1:  Standardized Mean Difference (SMD) of Variables After Propensity Score Matching**

| **Variable** | **Standardized Mean Difference** |
| --- | --- |
| Distance | 0.13 |
| **Age (years)** |  |
| < 65 | -0.12 |
| ≥ 65 | 0.12 |
| **Sex** |  |
| Female | 0.016 |
| Male | -0.016 |
| **Race/Ethnicity** |  |
| Non-Hispanic White | 0.023 |
| Other/race Ethnicity | -0.023 |
| **Obesity Status** |  |
| Not Obese | - 0.087 |
| Obese | 0.087 |
| **Immunocompromised Status** |  |
| Not Immunocompromise | -0.107 |
| Mild Immunocompromise | 0.078 |
| Moderate-Severe Immunocompromise | 0.062 |
| **Number of Other Comorbid Conditions** |  |
| None | -0.019 |
| One | -0.086 |
| Two or More | 0.104 |
| **Number of Vaccinations Prior to Infection** |  |
| 0 | -0.010 |
| 1 | -0.004 |
| 2 | -0.020 |
| ≥3 | 0.024 |
| **Insurance Status** |  |
| Private/Commercial/Medicare | 0.005 |
| Medicaid | -0.002 |
| Other | -0.005 |
| **Week** |  |
| 3/26 - 4/1 | 0.023 |
| 4/2 - 4/8 | 0.003 |
| 4/9 - 4/15 | 0.003 |
| 4/16 - 4/22 | -0.010 |
| 4/23 - 4/29 | 0.000 |
| 4/30 - 5/6 | -0.022 |
| 5/7 - 5/13 | 0.007 |
| 5/14 - 5/20 | 0.041 |
| 5/21 - 5/27 | -0.012 |
| 5/28 - 6/3 | -0.039 |
| 6/4 - 6/10 | -0.029 |
| 6/11 - 6/17 | 0.014 |
| 6/18 - 6/23 | 0.034 |

**Appendix Table 2: Immunocompromised Specifications**

| **Category** | **Medications** | **Conditions** |
| --- | --- | --- |
| Not Immunocompromised | No qualifying medications | No qualifying conditions |
| Mild  (Presence of either medication or condition if moderate/severe criteria not met) | - TNF-alpha inhibitors (infliximab, etanercept, golimumab, adalimumab) - Azathioprine alone - Mycophenolate alone - Systemic Prednisone - Systemic Methylprednisolone | - Elixhauser Rheumatic - Charlson or Elixhauser HIV without mention of AIDS |
| Moderate/Severe  (Presence of either medication or condition) | - Alemtuzumab - Azathioprine plus mTORi or calcineurin inhibitor - Belatacept - Eculizumab - Rituximab - Cyclophosphamide - Mycophenolate plus mTORi or calcineurin inhibitor - Thymoglobulin - Any Calcineurin inhibitor alone - Any mTORi alone - Any Chemotherapeutic Agent - Actinomycin - Alkylating Agent - Anthracenedione - Anthracycline - Anti-Metabolite - Anti-Microtubular - Aromatase Inhibitor - CDK Inhibitor - Cytotoxic - EZH2 Inhibitor - Hedgehog Inhibitor - Immunomodulatory Imide - Immunotherapy - Microtubule Inhibitor - mTOr Kinase Inhibitor - PI3K Inhibitor - Platinum-Based - Poly (ADP-Ribose) Polymerase Inhibitor - Proteasome Inhibitor - Targeted Monoclonal Antibody - Topoisomerase Inhibitor - Tyrosine Kinase Inhibitor - Vinca Alkaloid | - Charlson or Elixhauser HIV with AIDS based on CD4 count <200 cells/μL - Lymphoma - Cancer with Metastases - Tumor |

**Appendix Table 3: Baseline Characteristics by NMV/r Treatment Status for Full SARS-CoV-2 Positive Cohort, Prior to Propensity Matching**

| **Characteristic** | **NMV/r-Treated**  **n=4672** | **Untreated n=6542** |
| --- | --- | --- |
| **Age Group *** |  |  |
| 18-44 years | 1841 (39.4%) | 4437 (67.8%) |
| 45-64 years | 890 (19.0%) | 974 (14.9%) |
| ≥65 years | 1941 (41.5%) | 1131 (17.3%) |
| **Sex *** |  |  |
| Female | 2651 (56.7%) | 3760 (57.5%) |
| **Race/Ethnicity *** |  |  |
| Non-Hispanic White | 3638 (77.9%) | 4773 (73.0%) |
| Hispanic, any race | 381 (8.2%) | 750 (11.5%) |
| Non-Hispanic Black | 129 (2.8%) | 319 (4.9%) |
| Other | 179 (3.8%) | 340 (5.2%) |
| Missing | 345 (7.4%) | 360 (5.5%) |
| **Insurance Status *** |  |  |
| Private/Commercial | 2511 (53.7%) | 4609 (70.5%) |
| Medicare | 1704 (36.5%) | 1099 (16.8%) |
| Medicaid | 169 (3.6%) | 534 (8.2%) |
| None/Uninsured | 190 (4.1%) | 122 (1.9%) |
| Other/Unknown | 98 (2.1%) | 178 (2.7%) |
| **Immunocompromised *** |  |  |
| Mild | 525 (11.2%) | 424 (6.5%) |
| Moderate / Severe | 597 (12.8%) | 450 (6.9%) |
| **Obesity Status** |  |  |
| Yes | 1120 (24.0%) | 1065 (16.3%) |
| Missing | 243 (5.2%) | 242 (3.7%) |
| **Number of Other Comorbid Conditions *** |  |  |
| 0 | 1518 (32.5%) | 3731 (57.0%) |
| 1 | 1423 (30.5%) | 1473 (22.5%) |
| ≥2 | 1488 (31.8%) | 1096 (16.8%) |
| Missing | 243 (5.2%) | 242 (3.7%) |
| **Diabetes** |  |  |
| Yes | 701 (15.0%) | 500 (7.6%) |
| Missing | 243 (5.2%) | 242 (3.7%) |
| **Cardiovascular Disease** |  |  |
| Yes | 751 (16.1%) | 628 (9.6%) |
| Missing | 243 (5.2%) | 242 (3.7%) |
| **Pulmonary Disease** |  |  |
| Yes | 1325 (28.4%) | 1187 (18.1%) |
| Missing | 243 (5.2%) | 242 (3.7%) |
| **Renal Disease** |  |  |
| Yes | 317 (6.8%) | 273 (4.2%) |
| Missing | 243 (5.2%) | 242 (3.7%) |
| **Hypertension** |  |  |
| Yes | 1858 (39.8%) | 1418 (21.7%) |
| Missing | 243 (5.2%) | 242 (3.7%) |
| **Liver Disease** |  |  |
| Yes | 411 (8.8%) | 354 (5.4%) |
| Missing | 243 (5.2%) | 242 (3.7%) |
| **Number of Vaccinations *** |  |  |
| 0 | 809 (17.3%) | 1495 (22.9%) |
| 1 | 176 (3.8%) | 288 (4.4%) |
| 2 | 584 (12.5%) | 1163 (17.8%) |
| 3+ | 3103 (66.4%) | 3596 (55.0%) |
| **Week *** |  |  |
| 3/26 - 4/1 | 32 (0.7%) | 85 (1.3%) |
| 4/2 - 4/8 | 55 (1.2%) | 117 (1.8%) |
| 4/9 - 4/15 | 58 (1.2%) | 155 (2.4%) |
| 4/16 - 4/22 | 88 (1.9%) | 227 (3.5%) |
| 4/23 - 4/29 | 148 (3.2%) | 273 (4.2%) |
| 4/30 - 5/6 | 219 (4.7%) | 419 (6.4%) |
| 5/7 - 5/13 | 325 (7.0%) | 556 (8.5%) |
| 5/14 - 5/20 | 509 (10.9%) | 636 (9.7%) |
| 5/21 - 5/27 | 659 (14.1%) | 809 (12.4%) |
| 5/28 - 6/3 | 607 (13.0%) | 855 (13.1%) |
| 6/4 - 6/10 | 707 (15.1%) | 940 (14.4%) |
| 6/11 - 6/17 | 682 (14.6%) | 782 (12.0%) |
| 6/18 - 6/23 | 583 (12.5%) | 688 (10.5%) |

* Variables used for propensity matching; Abbreviations: NMV/r Nirmatrelvir-ritonavir

**Appendix Table 4: Baseline Characteristics by matching status for all NMV/r treated patients**

| **Characteristic** | **Matched**  **n=3614** | **Unmatched n=1058** |
| --- | --- | --- |
| **Age Group *** |  |  |
| 18-44 years | 1596 (44.2%) | 245 (23.2%) |
| 45-64 years | 755 (20.9%) | 135 (12.8%) |
| ≥65 years | 1263 (34.9%) | 678 (64.1%) |
| **Sex *** |  |  |
| Female | 2133 (59.0%) | 518 (49.0%) |
| **Race/Ethnicity *** |  |  |
| Non-Hispanic White | 3006 (83.2%) | 632 (59.7%) |
| Hispanic, any race | 351 (9.7%) | 30 (2.8%) |
| Non-Hispanic Black | 115 (3.2%) | 14 (1.3%) |
| Other | 142 (3.9%) | 37 (3.5%) |
| Missing | 0 (0.0%) | 345 (32.6%) |
| **Insurance Status *** |  |  |
| Private/Commercial | 2187 (60.5%) | 324 (30.6%) |
| Medicare | 1166 (32.3%) | 538 (50.9%) |
| Medicaid | 155 (4.3%) | 14 (1.3%) |
| None/Uninsured | 18 (0.5%) | 172 (16.3%) |
| Other/Unknown | 88 (2.4%) | 10 (0.9%) |
| **Immunocompromised *** |  |  |
| Mild | 454 (12.6%) | 71 (6.7%) |
| Moderate / Severe | 486 (13.4%) | 111 (10.5%) |
| **Obesity Status** |  |  |
| Yes | 1003 (27.8%) | 117 (11.1%) |
| Missing | 0 (0.0%) | 243 (23.0%) |
| **Number of Other Comorbid Conditions *** |  |  |
| 0 | 1374 (38.0%) | 144 (13.6%) |
| 1 | 1073 (29.7%) | 350 (33.1%) |
| ≥2 | 1167 (32.3%) | 321 (30.3%) |
| Missing | 0 (0.0%) | 243 (23.0%) |
| **Diabetes** |  |  |
| Yes | 555 (15.4%) | 146 (13.8%) |
| Missing | 0 (0.0%) | 243 (23.0%) |
| **Cardiovascular Disease** |  |  |
| Yes | 584 (16.2%) | 167 (15.8%) |
| Missing | 0 (0.0%) | 243 (23.0%) |
| **Pulmonary Disease** |  |  |
| Yes | 1063 (29.4%) | 262 (24.8%) |
| Missing | 0 (0.0%) | 243 (23.0%) |
| **Renal Disease** |  |  |
| Yes | 246 (6.8%) | 71 (6.7%) |
| Missing | 0 (0.0%) | 243 (23.0%) |
| **Hypertension** |  |  |
| Yes | 1377 (38.1%) | 481 (45.5%) |
| Missing | 0 (0.0%) | 243 (23.0%) |
| **Liver Disease** |  |  |
| Yes | 332 (9.2%) | 79 (7.5%) |
| Missing | 0 (0.0%) | 243 (23.0%) |
| **Number of Vaccinations *** |  |  |
| 0 | 654 (18.1%) | 155 (14.7%) |
| 1 | 142 (3.9%) | 34 (3.2%) |
| 2 | 488 (13.5%) | 96 (9.1%) |
| 3+ | 2330 (64.5%) | 773 (73.1%) |
| **Week *** |  |  |
| 3/26 - 4/1 | 24 (0.7%) | 8 (0.8%) |
| 4/2 - 4/8 | 43 (1.2%) | 12 (1.1%) |
| 4/9 - 4/15 | 45 (1.2%) | 13 (1.2%) |
| 4/16 - 4/22 | 71 (2.0%) | 17 (1.6%) |
| 4/23 - 4/29 | 111 (3.1%) | 37 (3.5%) |
| 4/30 - 5/6 | 171 (4.7%) | 48 (4.5%) |
| 5/7 - 5/13 | 269 (7.4%) | 56 (5.3%) |
| 5/14 - 5/20 | 410 (11.3%) | 99 (9.4%) |
| 5/21 - 5/27 | 491 (13.6%) | 168 (15.9%) |
| 5/28 - 6/3 | 451 (12.5%) | 156 (14.7%) |
| 6/4 - 6/10 | 546 (15.1%) | 161 (15.2%) |
| 6/11 - 6/17 | 521 (14.4%) | 161 (15.2%) |
| 6/18 - 6/23 | 461 (12.8%) | 122 (11.5%) |

* Variables used for propensity matching; Abbreviations: NMV/r Nirmatrelvir-ritonavir

**Appendix Table 5: Full Model Results for 28-day Hospitalization Primary Outcome**

| Characteristic | Adjusted OR | 95% CI | P-value |
| --- | --- | --- | --- |
| Treatment Status |  |  |  |
| Untreated | Reference |  |  |
| NMV/r-Treated | 0.48 | (0.31, 0.75) | <0.001 |
| Age Group |  |  |  |
| 18-44 | Reference |  |  |
| 45-64 | 0.54 | (0.25, 1.08) | 0.085 |
| ≥65 | 1.69 | (1.01, 2.85) | 0.046 |
| Sex |  |  |  |
| Female | Reference |  |  |
| Male | 1.33 | (0.88, 2.01) | 0.18 |
| Race/Ethnicity |  |  |  |
| Non-Hispanic White | Reference |  |  |
| Hispanic | 0.83 | (0.39, 1.60) | 0.598 |
| Non-Hispanic Black | 0.92 | (0.33, 2.06) | 0.856 |
| Other | 0.66 | (0.14, 1.94) | 0.494 |
| Insurance Status |  |  |  |
| Private/Commercial/Medicare | Reference |  |  |
| Medicaid | 3.45 | (1.72, 6.58) | <0.001 |
| Other (None/Uninsured/Unknown) | 1.29 | (0.26, 3.92) | 0.714 |
| Obesity Status |  |  |  |
| No | Reference |  |  |
| Yes | 1.71 | (1.10, 2.64) | 0.018 |
| Immunocompromised Status |  |  |  |
| No | Reference |  |  |
| Mild | 1.7 | (0.90, 3.04) | 0.101 |
| Moderate/Severe | 2.51 | (1.49, 4.22) | <0.001 |
| Number of Comorbid Conditions |  |  |  |
| 0 | Reference |  |  |
| 1 | 1.69 | (0.87, 3.38) | 0.124 |
| ≥2 | 4.09 | (2.16, 8.05) | <0.001 |
| Vaccination Status (# of doses) |  |  |  |
| 0 | Reference |  |  |
| 1 | 0.83 | (0.26, 2.11) | 0.721 |
| 2 | 0.92 | (0.49, 1.68) | 0.78 |
| ≥3 | 0.51 | (0.31, 0.86) | 0.012 |
| Time |  |  |  |
| One-week increase | 1.04 | (0.96, 1.12) | 0.343 |
